# Evaluating Accuracy and Reproducibility of Large Language Model Performance in Pharmacy Education

**DOI:** 10.1101/2024.03.21.24304667

**Authors:** Amoreena Most, Mengxuan Hu, Huibo Yang, Tianming Liu, Xianyan Chen, Sheng Li, Steven Xu, Zhengliang Liu, Andrea Sikora

**Affiliations:** University of Georgia College of Pharmacy, Augusta, GA, USA; School of Data Science, University of Virginia, Charlottesville, VA; Department of Computer Science, University of Georgia, Athens, GA; University of Georgia Statistical Consulting Center, Athens, GA; University of Georgia College of Pharmacy, Department of Clinical and Administrative Pharmacy, Augusta, GA, USA

**Keywords:** Large language model, artificial intelligence, pharmacy

## Abstract

The purpose of this study was to compare performance of ChatGPT (GPT-3.5), ChatGPT (GPT-4), Claude2, Llama2-7b, and Llama2-13b on 219 multiple-choice questions focusing on critical care pharmacotherapy. To further assess the ability of engineering LLMs to improve reasoning abilities and performance, we examined responses with a zero-shot Chain-of-Thought (CoT) approach, CoT prompting, and a custom built GPT (PharmacyGPT). A 219 multiple-choice questions focused on critical care pharmacotherapy topics used in Doctor of Pharmacy curricula from two accredited colleges of pharmacy was compiled for this study. A total of five LLMs were evaluated: ChatGPT (GPT-3.5), ChatGPT (GPT-4), Claude2, Llama2-7b, and Llama2-13b. The primary outcome was response accuracy. Of the five LLMs tested, GPT-4 showed the highest average accuracy rate at 71.6%. A larger variance indicates lower consistency and reduced confidence in its answers. Llama2-13b had the lowest variance (0.070) of all the LLMs, but performed with an accuracy of 41.5%. Following analaysis of overall accuracy, performance on knowledge- vs. skill-based questions were assessed. All five LLMs demonstrated higher accuracy on knowledge-based questions compared to skill-based questions. GPT-4 had the highest accuracy for knowledge- and skill-based questions, with an accuracy of 87% and 67%, respectively. Response accuracy from LLMs in the domain of clinical pharmacy can be improved by using prompt engineering techniques.

## Introduction

Large language models (LLMs) have shown remarkable abilities in the medical domain, including passing medical licensure exams, diagnosing disease states, and clinical decision making; however, these task have largely focused on structured diagnostic problems and have limited pharmacy domain testing. level.^1–3^ ^4^ Within the field of clinical pharmacy, the performance of LLMs have been tested for deprescribing benzodiazepines, identifying drug-herb interactions, and performance on a national pharmacist examination, showing early promise.^5–8^ Each year it is estimated over 6.3 billion prescription medications are dispensed and over 7 million patients will experience a medication error. Given the complexity of medication data and ability of LLMs to process large datasets, they may serve as an important tool towards making medication use safer.

**Table 1.** Statement of Significance.

|  |  |
|---|---|
| **Problem** | Large Language Models (LLMs) reasoning processes for clinical pharmacy decision making has not been explored and remains unknown. |
| **What is already known** | LLMs have demonstrated promise in pharmacy tasks but require further investigation to improve performance in the clinical pharmacy domain. |
| **What this paper adds** | This paper demonstrates LLM performance on clinical pharmacy questions can be improved with prompt engineering. |

However, most LLMs are trained on data from widely available corpus (e.g., the Internet), which creates the potential for problems in domains marked by highly technical language.^9^ Moreover, deconstructing LLMs reasoning abilities have been identified as a significant challenge.^10,11^ There have been calls for thoughtful evaluation and regulation of artificial intelligence prior to implementation in the healthcare setting.^12^ Approaches for understanding LLMs reasoning processes to improve performance include fine-tuning a pre-trained LLM or building a LLM with a custom dataset.^13^ Limited studies have rigorously explored strategies to benchmark and improve LLM performance in the medication decision-making domain.

The purpose of this study was to compare performance of ChatGPT (GPT-3.5), ChatGPT (GPT-4), Claude2, Llama2-7b, and Llama2-13b on 219 multiple-choice questions focusing on critical care pharmacotherapy. To further assess the ability of engineering LLMs to improve reasoning abilities and performance, we examined responses with a zero-shot Chain-of-Thought (CoT) approach, CoT prompting, and a custom built GPT (PharmacyGPT).

**Figure 1.**
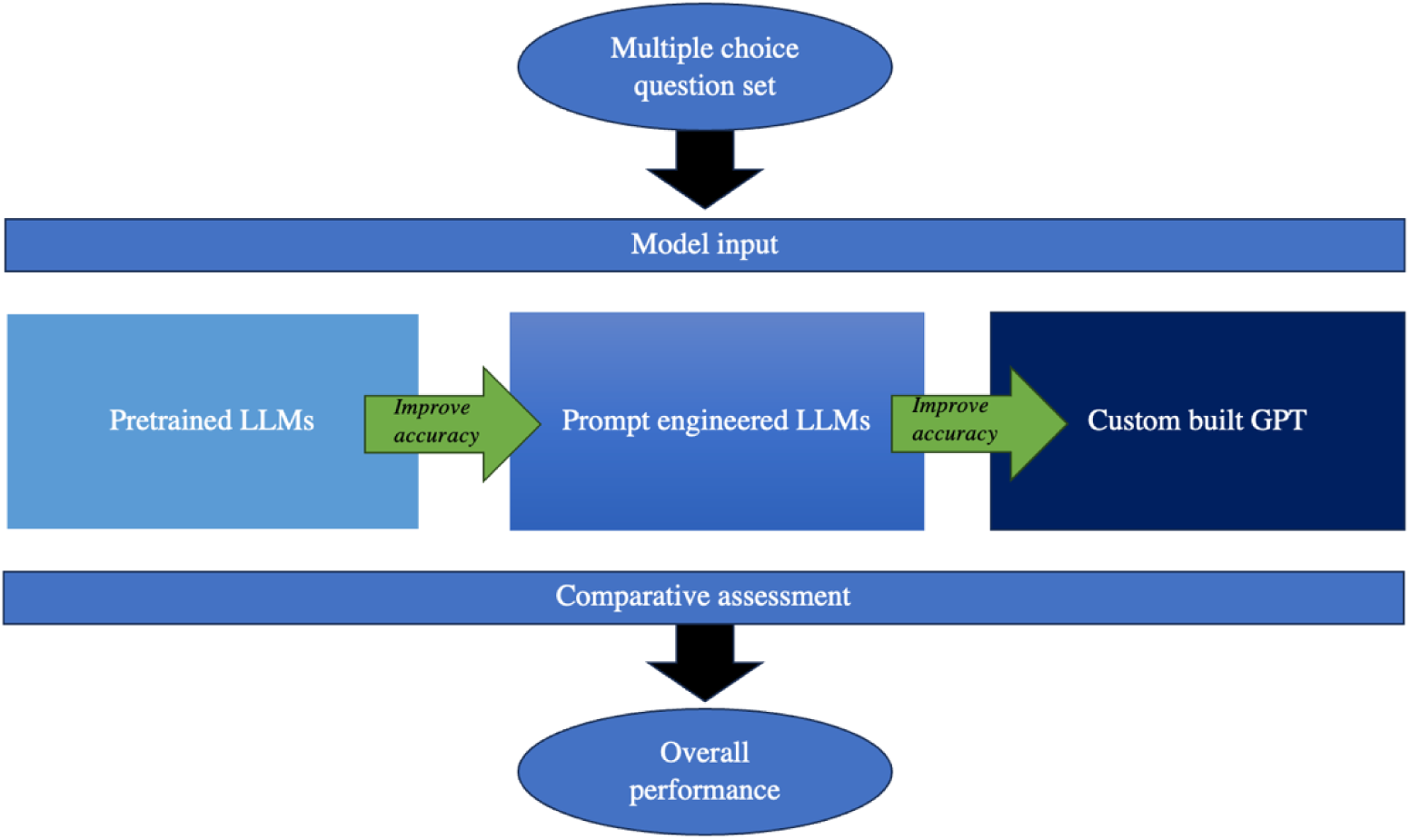
Study methodology

**Figure 2.**
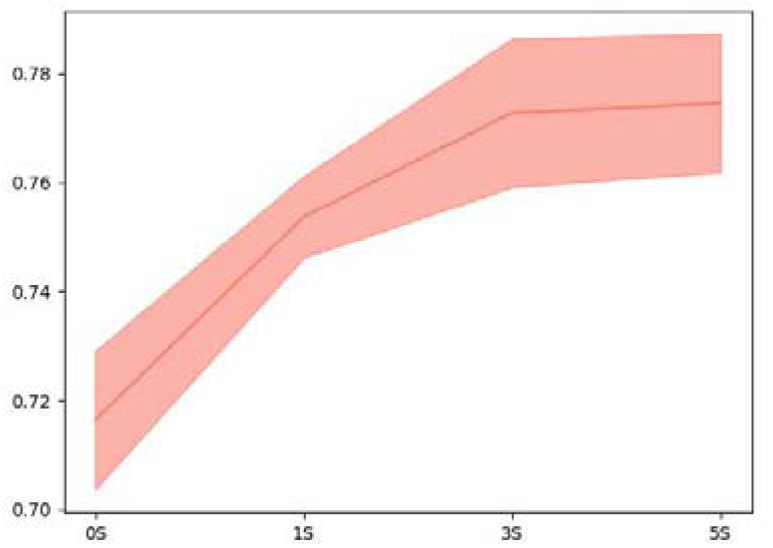
Response accuracy across few shot CoT

**Figure 3.**
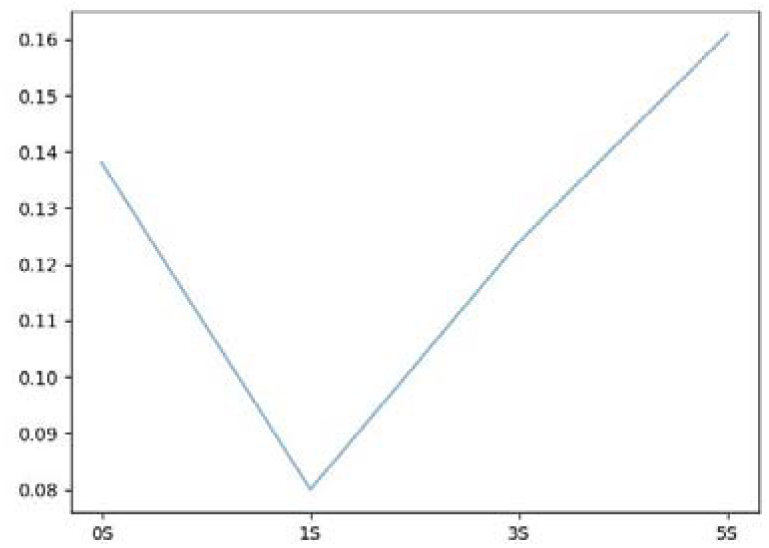
Response variance across few shot CoT

**Figure 4.**
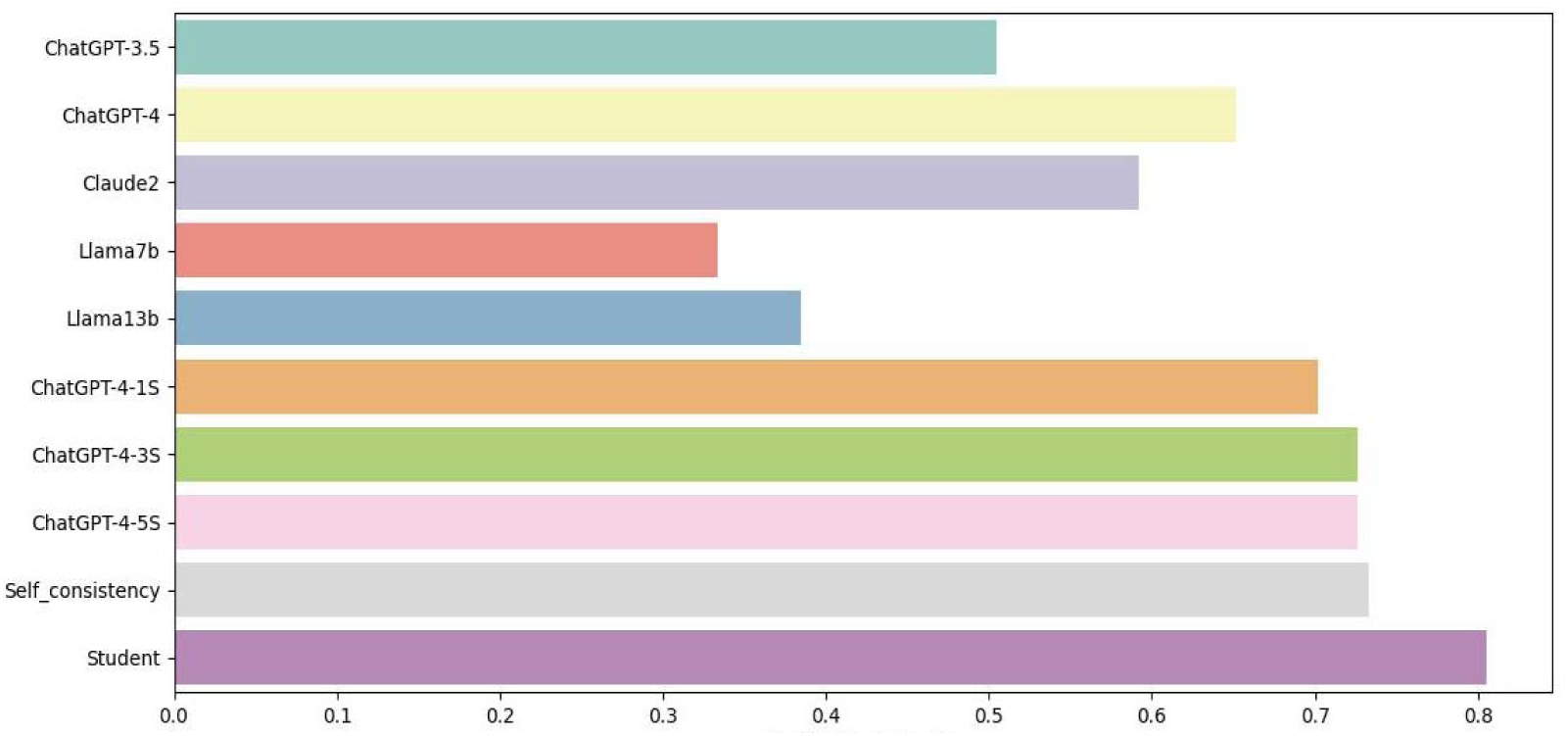
Response accuracy across LLMs and students

**Figure 5.**
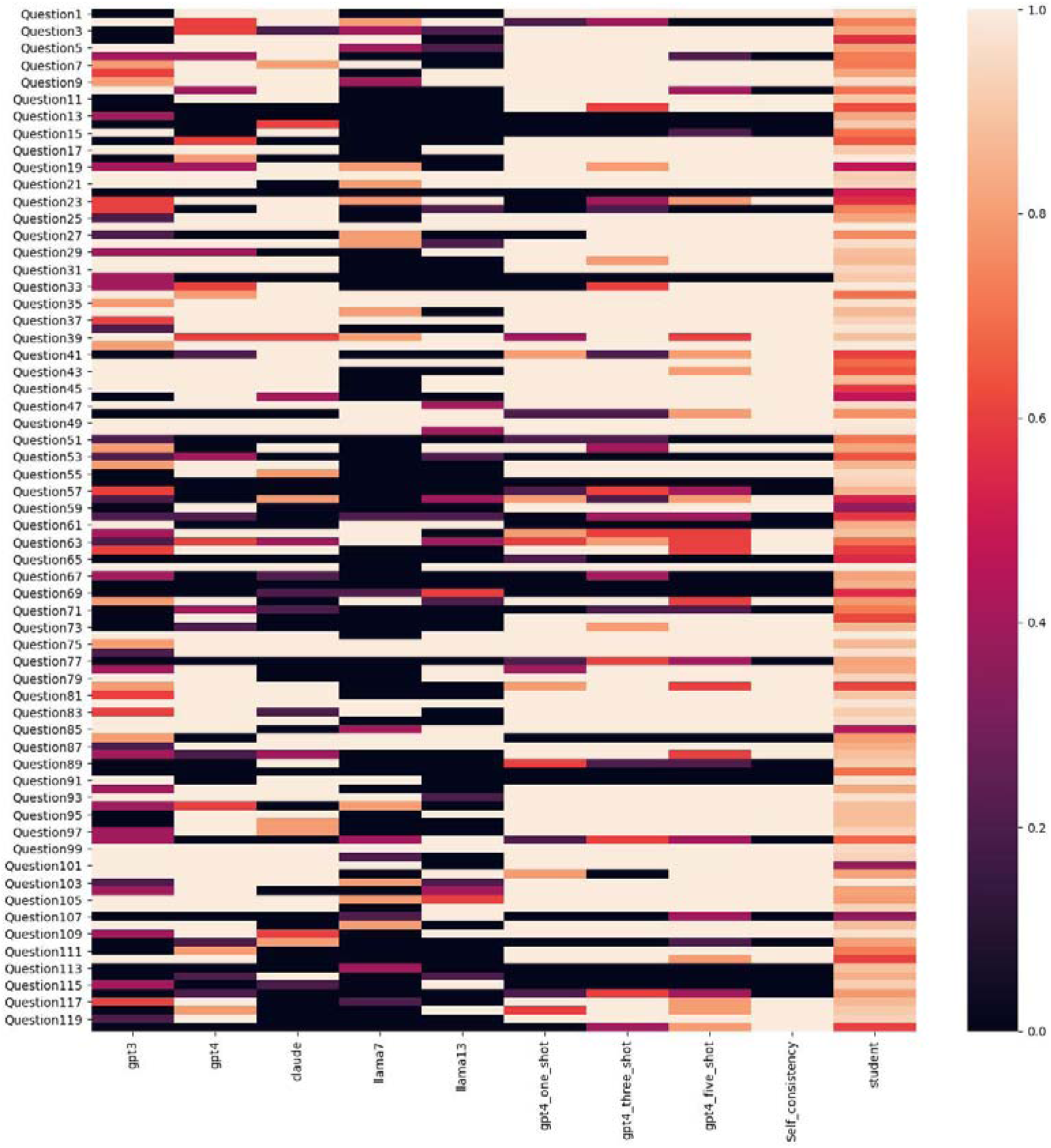
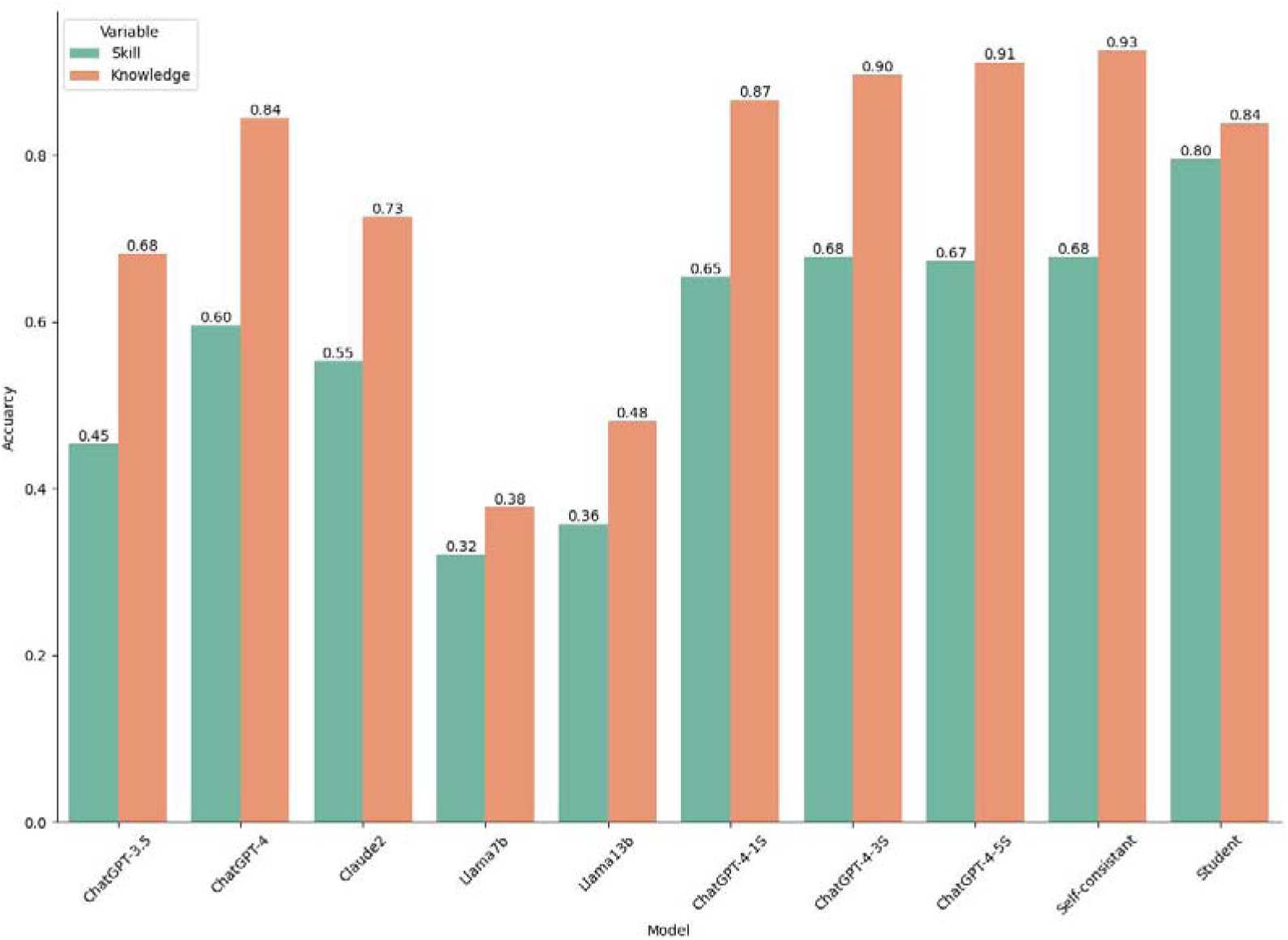
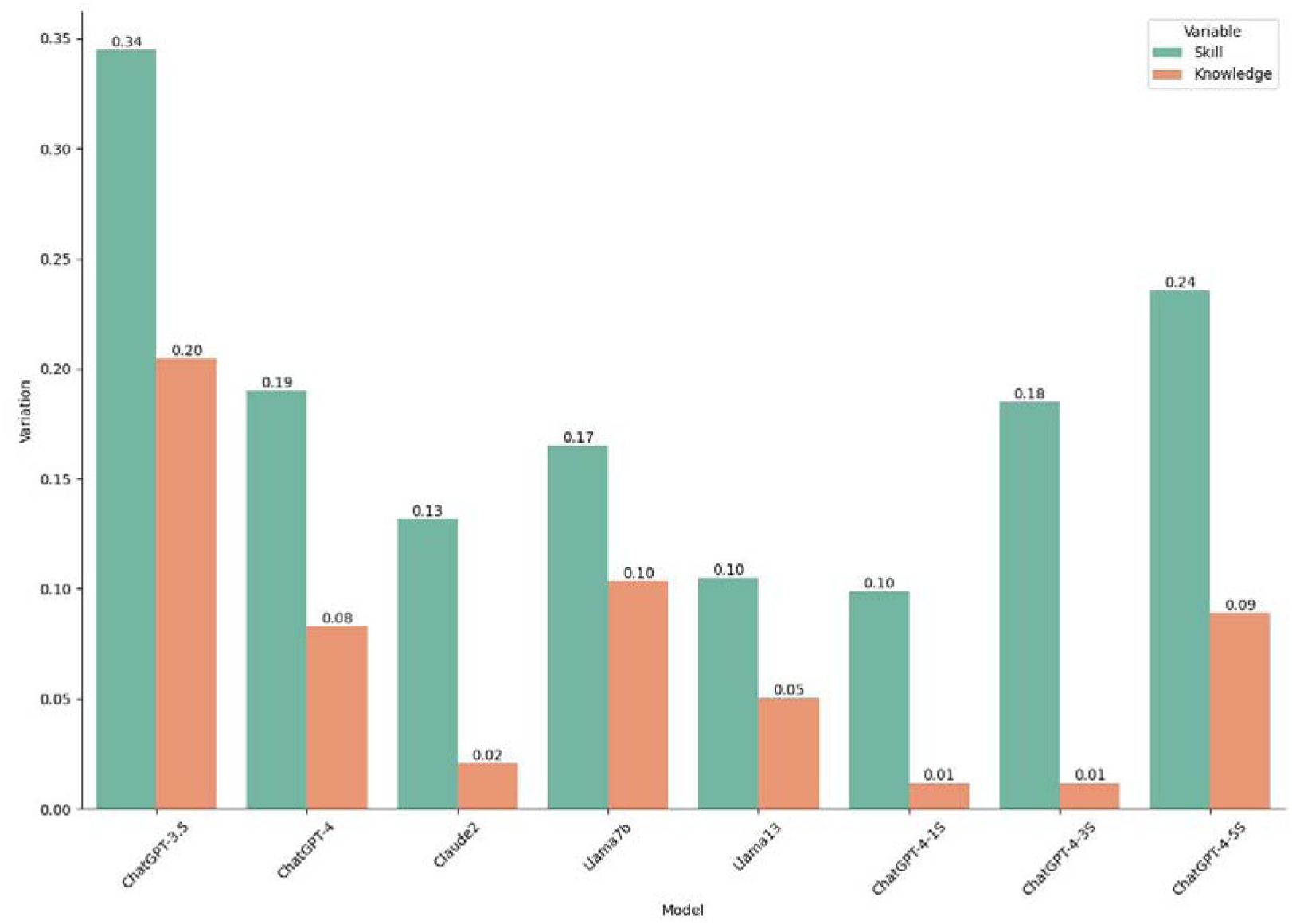

**Figure 6.**
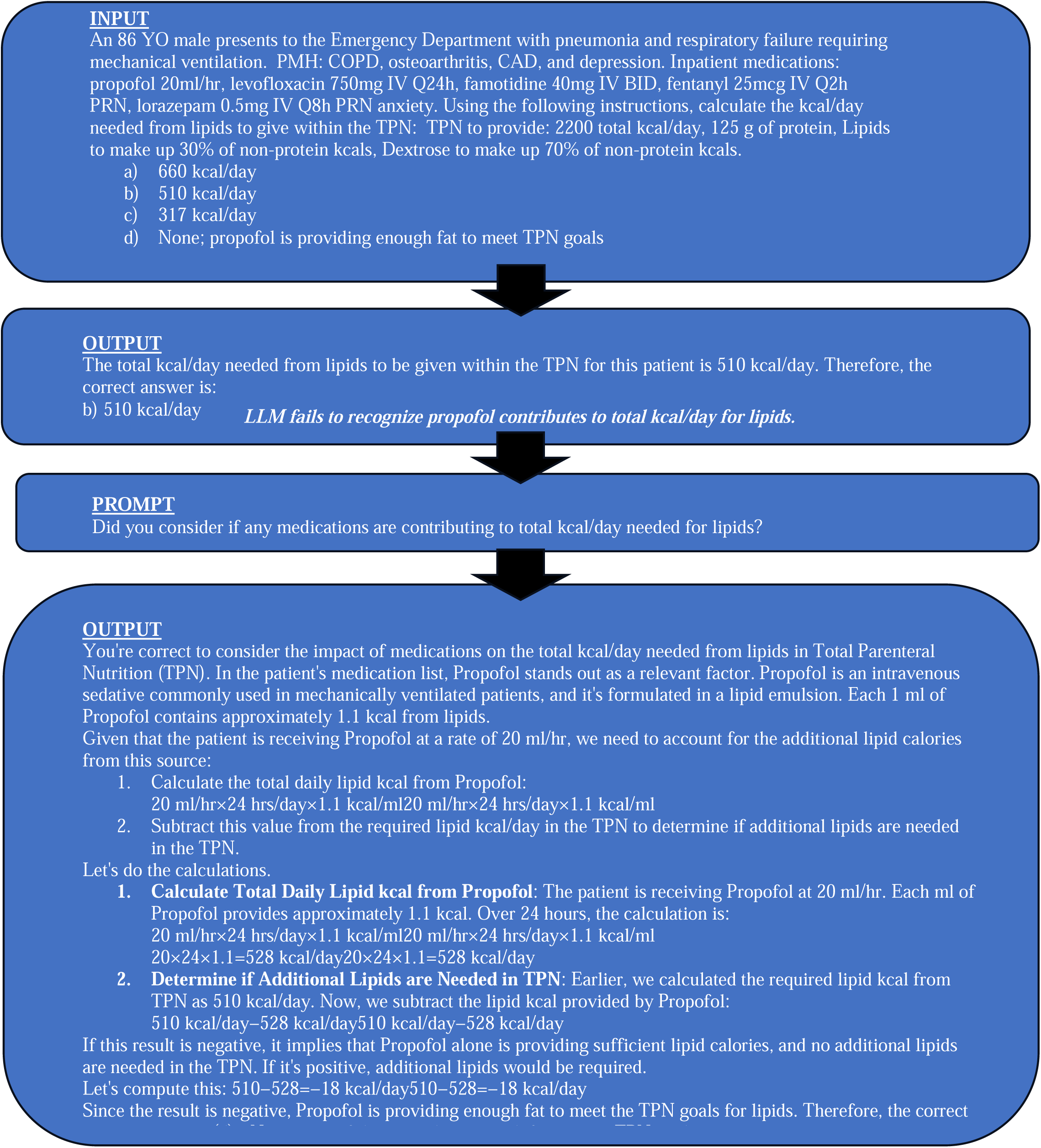
Prompt engineering example

## Methods

### Data source

A 219 multiple-choice questions focused on critical care pharmacotherapy topics used in Doctor of Pharmacy curricula from two accredited colleges of pharmacy was compiled for this study. Questions were written for students in their third-year pharmacy school who participated in critical care elective and pharmacotherapy course. Questions on the following topics were assesed: metabolic disorders (26 questions), pain/agitation/delirium (23 questions), respiratory disorders (22 questions), toxicology (20 questions), hemodynamics (17 questions), acid-base (16 questions), neurologic emergencies (14 questions), gastrointestinal disorders (10 questions), prophylaxis (10 questions), advanced cardiac life support (9 questions), nutrition (9 questions), renal (9 questions), sedation management (9 questions), fluids (7 questions), anticoagulation reversal (4). Of the 219 questions, 27 required calculations based on patient specific parameters (e.g., weight, renal function, laboratory parameters). Questions were formatted to have four answer choices and images were converted to textual input. Additionally, questions were further categorized into knowledge- or skill-based, with knowledge questions testing fact recall and skill testing application of pharmacy knowledge to simple patient cases.

### Study design

A total of five LLMs were evaluated: ChatGPT (GPT-3.5), ChatGPT (GPT-4), Claude2, Llama2-7b, and Llama2-13b. The primary outcome was response accuracy. Secondary outcomes included response variance and comparison to student performance. To assess consistency of response, questions were inputted into each LLM five separate times and numeric values were assigned (1, 2, 3, 4) to the four answer choices in each question. Variance was calculated from the response accuracy for each individual LLM after the five runs.

### Initialization prompt

Input was standardized to generate output that provided correct answers and explanations. The following system prompt was utilized: “This is a midterm exam for the critical care elective course in pharmacy school. Please select the most correct answer from the following multiple-choice options and give your reason why you chose it. Please follow the following format to answer the question: The correct answer is ---. The reason is ---.”

### Zero-Shot Chain-of-Thought

A Zero-Shot chain-of-thought (CoT) approach was then employed by including “Let’s think step by step” in the prompt and requesting the model to output the answer along with the corresponding explanation directly. Zero-Shot CoT was applied to each of the five LLMs and was evaluated in five separate trials. The Zero-Shot CoT LLM answers were compared to the pretrained LLM to assess if there was improvement in accuracy or variance.

### Few-Shot Chain-of-Thought

Due to the complex reasoning required to answer skill-based clinical pharmacy questions, five chain-of-thought (CoT) prompts were created to improve LLM accuracy of responses. CoT was applied to GPT4 and was evaluated in five separate trials. The CoT prompted GPT-4 answers were compared with the pretrained GPT4 results and Zero-Shot CoT GPT-4 results to assess if there was improvement in accuracy or variance. A full overview of the CoT prompts created and applied are available in the supplemental materials.

### Customized GPT

ChatGPT-4 offers the ability for a user to create a customizable GPT. We built a ChatGPT based on relevant pharmacy course materials as a proof of concept to improve GPT-4 accuracy and reproducibility. These results were then compared to the pretrained non-CoT prompted GPT-4 results and CoT prompted GPT-4 results.

### LLMs to students

Student performance was available for 120 multiple choice questions. Response accuracy and variance on knowledge- and skill-based questions from the unprompted LLMs (ChatGPT (GPT-3.5), ChatGPT (GPT-4), Claude2, Llama2-7b, and Llama2-13b) and GPT-4 engineered with few-shot CoT were assessed for the 120 questions and then compared to student performance.

## Results

### Initialization prompt

Table 1 shows the performance of five LLMs: ChatGPT (GPT-3.5), ChatGPT (GPT-4), Claude2, Llama2-7b, and Llama2-13b accuracy from individual runs and calculated variance after the five runs. Of the five LLMs tested, GPT-4 showed the highest average accuracy rate at 71.6%. A larger variance indicates lower consistency and reduced confidence in its answers. Llama2-13b had the lowest variance (0.070) of all the LLMs, but performed with an accuracy of 41.5%. Following analaysis of overall accuracy, performance on knowledge- vs. skill-based questions were assessed. All five LLMs demonstrated higher accuracy on knowledge-based questions compared to skill-based questions. GPT-4 had the highest accuracy for knowledge- and skill-based questions, with an accuracy of 87% and 67%, respectively.

**Table 1.** Response accuracy and variance of LLMs.

| LLM | Accuracy-Run1 | Accuracy-Run2 | Accuracy-Run3 | Accuracy-Run4 | Accuracy-Run5 | Accuracy-ALL | Variance-ALL |
|---|---|---|---|---|---|---|---|
| ChatGPT-3.5 | 0.559 | 0.536 | 0.518 | 0.550 | 0.545 | 0.542 | 0.304 |
| ChatGPT-4 | 0.732 | 0.705 | 0.714 | 0.727 | 0.705 | 0.716 | 0.135 |
| Claude2 | 0.609 | 0.609 | 0.623 | 0.618 | 0.618 | 0.615 | 0.090 |
| Llama2-7b | 0.364 | 0.382 | 0.364 | 0.350 | 0.355 | 0.3627 | 0.210 |
| Llama2-13b | 0.400 | 0.400 | 0.445 | 0.418 | 0.414 | 0.415 | 0.070 |

**Table 2.** Response accuracy and variance of LLMs answering skill vs. knowledge based questions.

| LLM | Accuracy-knowledge | Accuracy-skill | Variance-knowledge | Variance-skill |
|---|---|---|---|---|
| ChatGPT-3.5 | 0.69 | 0.50 | 0.22 | 0.33 |
| ChatGPT-4 | 0.87 | 0.67 | 0.08 | 0.15 |
| Claude2 | 0.75 | 0.57 | 0.88 | 0.09 |
| Llama2-13b | 0.51 | 0.39 | 0.06 | 0.07 |

### Prompt engineering performance

Table 3 presents the response accuracy and variance with a zero-shot CoT approach. All five LLMs performed similiarily with a zero-shot COT approach compared to the original initialization prompt used. GPT-4 outperformed the other models with an average accuracy rate at 71.6%, while Llama2-7b had the lowest average accuracy rate at 34.5%. As more CoT examples were inputted into the model, accuracy improved while variance increased.

**Table 3.** Response accuracy and variance of LLMs with zero-shot CoT.

| LLM | Accuracy-Run1 | Accuracy-Run2 | Accuracy-Run3 | Accuracy-Run4 | Accuracy-Run5 | Accuracy-ALL | Variance-ALL |
|---|---|---|---|---|---|---|---|
| ChatGPT-3.5 | 0.550 | 0.536 | 0.532 | 0.568 | 0.541 | 0.545 | 0.329 |
| ChatGPT-4 | 0.731 | 0.705 | 0.714 | 0.727 | 0.705 | 0.716 | 0.138 |
| Claude2 | 0.595 | 0.595 | 0.618 | 0.600 | 0.605 | 0.603 | 0.080 |
| Llama2-7b | 0.350 | 0.3455 | 0.336 | 0.336 | 0.355 | 0.345 | 0.137 |
| Llama2-13b | 0.386 | 0.423 | 0.405 | 0.423 | 0.414 | 0.410 | 0.095 |

**Table 4.**
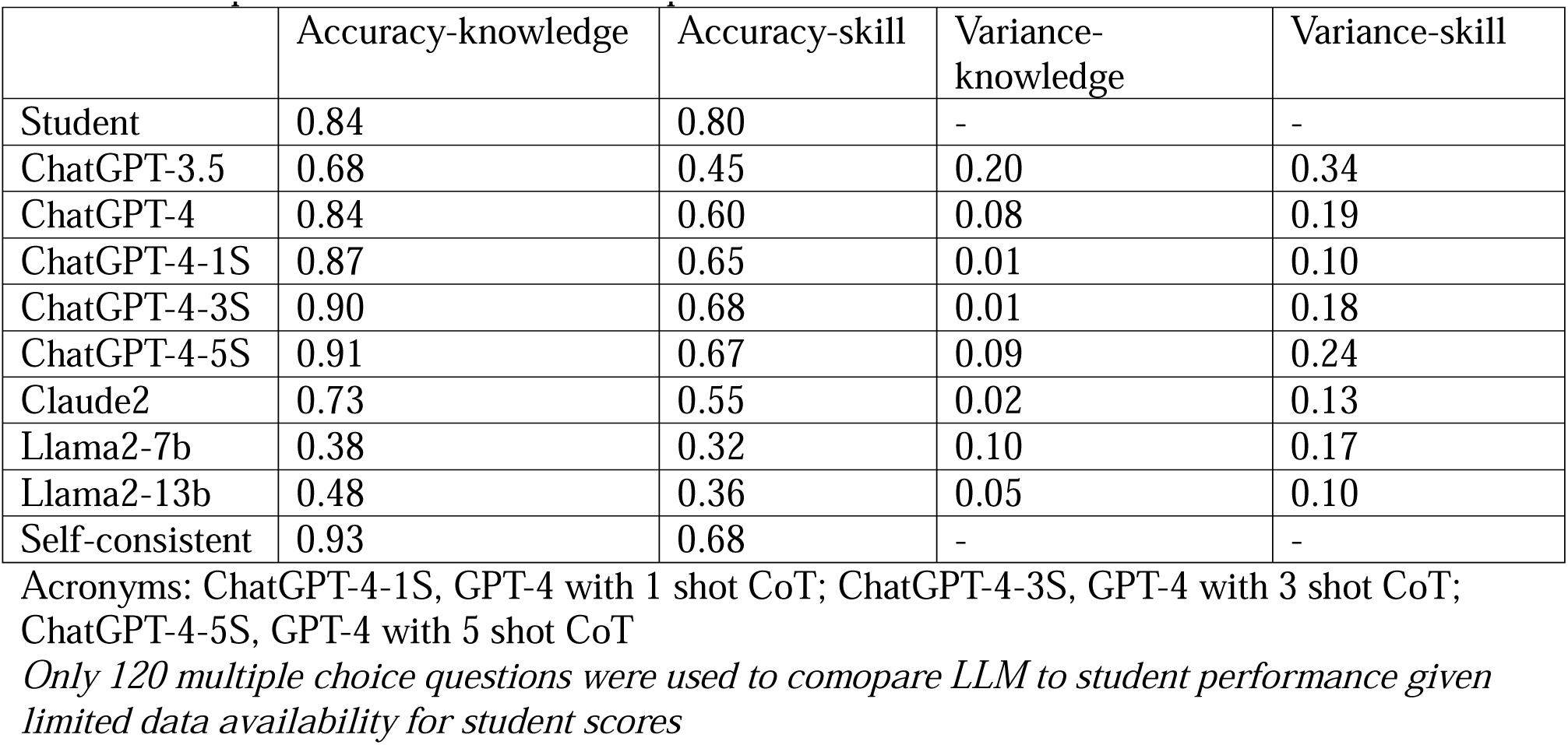
Comparison of LLMs to student performance.

### LLMs to students

GPT-4 with 5 shot CoT had the highest accuracy for knowledge-based questions, and outperformed the student average in this domain (91% vs. 84%). Accuracy for both knowledge-and skill based questions improved as additional CoT examples were provided; however, the models performance even with the highest accuracy for skill-based questions was lower than the student average (68% vs. 80%).

## Discussion

In this study, we demonstrate response accuracy of LLMs in the clinical pharmacy domain can be improved through specific prompt engineering techniques. Among the five LLMs assessed in this study (GPT-3.5, GPT-4, Claude2, Llama2-7b, and Llama2-13b), GPT-4 consistently displayed the highest response accuracy when multiple prompt techniques were employed. When questions were entered without utilizing prompt-engineering techniques, GPT-4 had a response accuracy for knowledge-based questions similar to a third year pharmacy student (84%). Use of CoT and self-consistency prompting increased GPT-4 response accuracy to outperform pharmacy students in the domain of knowledge-based pharmacy questions (91% and 93%, respectfully). These prompt engineering techniques showed minimal improvement in response accuracy for skill-based questions. To our knowledge, this is the first study to examine multiple prompt engineering techniques to improve LLM performance in the field of pharmacy.

Although large language models have demonstrated remarkable success across a wide spectrum of natural language processing (NLP) tasks, their reasoning abilities have been identified as a significant challenge.^10,11^ To address this issue, one intuitive approach is to either train a model from scratch using a dataset augmented with rationales or fine-tune a pre-trained large language model.^14^ In-context learning draws inspiration from human reasoning patterns when encountering a new task. A concise task instruction (e.g., “Please help me add the last two numbers in an array together and return the result.”) or a few related examples (e.g., “For an array [2,5,6,8], the result is 14.”) are often sufficient for humans to successfully complete the task to a satisfactory degree.^15^ Recent research endeavors have proposed in-context learning strategies, such as zero-shot CoT and CoT prompting, to enhance the reasoning capacity of LLMs.^15,16^

Previous research has demonstrated LLMs are capable of zero-shot reasoning, suggesting that a simple zero-shot prompt such as “Let’s think step by step” after each query can guide LLMs to answer questions in a CoT manner.^17^ The zero-shot CoT approach is an efficient method to model training as it eliminates the need for manually crafting intricate task-specific prompts for different tasks. In this study, model performance was similar with the original initialization prompt and when a zero-shot CoT approach was employed. Given our initialization prompt asked for reasoning to be provided with the output, it likely served as a modified zero-shot CoT approach and can explain why minimal difference was seen. CoT prompting provides carefully designed CoT examples to the LLM, allowing the LLMs to decompose a complex reasoning query into multiple steps and solve them step by step. Our study demonstrated that CoT prompting can improve LLM response. As additional CoT prompts were inputted into the LLM, performance improved in a linear fashion. Further research should focus on strategies to optimize CoT prompting examples and improve LLM performance.

In our study, all LLMs tested consistently demonstrated higher accuracy in answering knowledge-based questions when compared to skill-based questions. Knowledge-based questions are well-defined and widely accessible in textbooks and online resources, which LLMs have been trained on. In contrast, skill-based questions require reasoning abilities. Significant barriers to integrating LLMs into the healtcare system exist, including _xyz. Previous studes have demonstrated performance of ChatGPT on medical exams varies depending upon the specialty, with ChatGPT achieving a passing grade on Neurosurgery board finals yet failing a gastroenterology board-like examination.^18^ As the potential of LLMs to serve as a clinical decision support tool in the medical field continues to evolve, further research is needed to address the current limitations of their clinical reasoning capabilities.

## Conclusion

Response accuracy from LLMs in the domain of clinical pharmacy can be improved by using prompt engineering techniques. LLMs have demonstrated potential to serve as clinical pharmacy decision making tool. Future research is needed to optimize prompt engineering strategies and improve clinical pharmacy reasoning capabilities of LLMs.

## Data Availability

All data produced in the present study are available upon reasonable request to the authors

## Acknowledgements

The authors acknowledge William Hsieh for assistance with creating figures for this article.

## Supplemental

Chain-of-thought examples provided to LLMs:

1. Q: A 62-year-old male (70 kg) with no significant past medical history is admitted to the Medical ICU for acute hypoxic respiratory failure secondary to hospital acquired pneumonia. Current CrCl = 20 mL/min. Vancomycin and piperacillin/tazobactam are started for empiric antimicrobial coverage. How much vancomycin should be administered for the initial dose?

a) 1000 mg

b) 1250 mg

c) 1750 mg

d) 2250 mg

A: The recommended loading dose of vancomycin is 25 mg/kg in patients who are critically ill, regardless of renal function. 70 kg x 25 mg/kg = 1750 mg. The initial dose of vancomycin should be 1750 mg, therefore the correct answer is C.

2. Q: A 91-year-old female (80 kg, 5’2”) presents to the Emergency Department from a skilled nursing facility with altered mental status. Initial vitals: BP 81/43, HR 135, RR 24, Temp 102.1 (F). How much fluid should the patient initially receive for suspected sepsis?

a) 1250 mL

b) 1500 mL

c) 1750 mL

d) 2000 mL

A: The recommended minimum amount of crystalloid fluid resuscitation for a patient presenting with sepsis is 30 mL/kg based on ideal body weight. The patient’s ideal body weight is 50 kg. 30 mL/kg x 50 kg = 1500 mL. The initial amount of fluid the patient should receive is 1500 mL, therefore the correct answer is B.

3. Q: A 53-year-old male (110kg) is on hour 52 of admission to the Surgical ICU after an emergent exploratory laparotomy. Throughout his admission he has received 2 liters of IV fluids, started on TPN at 42 mL/hr (has received for 24 hours), has had a urine output of 0.5 cc/kg/hr, and 500 mL output from his nasogastric tube. What is the patient’s net fluid balance?

a) -322 mL

b) -332 mL

c) -342 mL

d) -352 mL

A: The patient’s total fluid intake is the sum of the IV fluids he received from IV fluids and TPN. He has received TPN at 42 mL/hr for 24 hours. 42 x 24 = 1008 mL. His total fluid intake is 3008 mL when adding up IV fluids and TPN fluids. The patient’s total fluid output is the sum of urine and nasogastric output. We will assume his urine output has remained the same throughout the 52 hours of admission to the surgical ICU. 0.5 cc/kg/hr x 110 kg x 52 hours = 2860 mL. His total fluid output is 3360 when adding up urine and nasogastric output. Net fluid balance = Total intake – Total output. 3008-3360 = -352 mL, therefore the correct answer is D.

4. Q: A 68 year-old-female (92 kg) is admitted to the Cardiac ICU for cardiogenic shock. PMH: HFrEF (EF 25%), HLD, anxiety, PE. She is currently mechanically ventilated. Medications: Nitroprusside 0.5 mcg/kg/min, furosemide 80 mg/hr, IV famotidine 20 mg BID, midazolam 4 mg/hr, fentanyl 100 mcg/hr, heparin 18u/kg/hr. Current CPOT: 1, RASS -4. Based on this information, how should her sedation be managed?

a) Do not adjust fentanyl infusion, decrease midazolam infusion. Target RASS of -2 to +1

b) Increase fentanyl infusion, do not adjust midazolam infusion. Target RASS of -5

c) Do not adjust fentanyl infusion, decrease midazolam infusion. Target RASS of -3 to - 2

d) Decrease fentanyl infusion, do not adjust midazolam infusion. Target RASS of +2 to +3

A: Light sedation is recommended for critically ill, mechanically ventilated adults unless deep sedation is required (ie, when neuromuscular blockade is indicated). Light sedation is defined as a RASS of -2 to +1. The patient’s current RASS is -4, which indicates she is too heavily sedated. Therefore, her fentanyl infusion or midazolam infusion needs to be decreased. Since her current CPOT is 1 indicating her pain is likely minimal or not present, her fentanyl infusion should not be adjusted. Her midazolam infusion should be decreased until a RASS of -2 to +1 is reached, therefore the correct answer is A.

5. Q: A 75-year-old female (65 kg, 5’4”) is admitted to the Neuro ICU for status epilepticus. Home medications: Aspirin 81 mg, atorvastatin 80 mg daily, sertraline 100 mg daily, phenytoin 100 mg TID. Phenytoin level upon arrival to ICU: 7 mcg/mL. How much IV phenytoin should be administered to achieve a target level of 18 mcg/mL?

a) 450 mg

b) 475 mg

c) 500 mg

d) 525 mg

A: Standard volume of distribution (Vd) of phenytoin is 0.7 L/kg. Vd = 0.7L/kg x 65 kg = 45.5 L. Dose (mg) = Vd x (Desired concentration – Measure concentration). 45.5 x (18-7) = 500.5 mg. The patient should receive IV phenytoin 500 mg (rounded to nearest tenth) for a desired concentration of 18, therefore the correct answer is C.

